# Differential overlap in human and animal fecal microbiomes and resistomes in rural versus urban Bangladesh

**DOI:** 10.1101/2021.05.13.21257188

**Authors:** Jenna M. Swarthout, Erica R. Fuhrmeister, Latifah Hamzah, Angela R. Harris, Mir A. Ahmed, Emily S. Gurley, Syed M. Satter, Alexandria B. Boehm, Amy J. Pickering

## Abstract

**Background:** Low- and middle-income countries (LMICs) bear the largest mortality burden due to antimicrobial-resistant infections. Small-scale animal production and free-roaming domestic animals are common in many LMICs, yet data on zoonotic exchange of gut bacteria and antimicrobial resistance genes (ARGs) in low-income communities are sparse. Differences between rural and urban communities in population density, antibiotic use, and cohabitation with animals likely influence the frequency of transmission of gut bacterial communities and ARGs between humans and animals. Here, we determined the similarity in gut microbiomes, using 16S rRNA gene amplicon sequencing, and resistomes, using long-read metagenomics, between humans, chickens, and goats in rural compared to urban Bangladesh.

**Results:** Gut microbiomes were more similar between humans and chickens in rural (where cohabitation is more common) compared to urban areas, but there was no difference for humans and goats. Urbanicity did not impact the similarity of human and animal resistomes; however, ARG abundance was higher in urban animals compared to rural animals. We identified substantial overlap of ARG alleles in humans and animals in both settings. Humans and chickens had more overlapping ARG alleles than humans and goats. All fecal hosts carried ARGs on contigs classified as potentially pathogenic bacteria – including *Escherichia coli, Campylobacter jejuni, Clostridiodes difficile*, and *Klebsiella pneumoniae*.

**Conclusions:** While the development of antimicrobial resistance in animal gut microbiomes and subsequent transmission to humans has been demonstrated in intensive farming environments and high-income countries, evidence of zoonotic exchange of antimicrobial resistance in LMIC communities is lacking. This research provides genomic evidence of overlap of antimicrobial resistance genes between humans and animals, especially in urban communities, and highlights chickens as important reservoirs of antimicrobial resistance. Chicken and human gut microbiomes were more similar in rural Bangladesh, where cohabitation is more common. Incorporation of long-read metagenomics enabled characterization of bacterial hosts of resistance genes, which has not been possible in previous culture-independent studies using only short-read sequencing. These findings highlight the importance of developing strategies for combatting antimicrobial resistance that account for chickens being reservoirs of ARGs in community environments, especially in urban areas.

## Background

In low- and middle-income countries (LMICs), animal husbandry is an important source of household income and nutrition [1, 2]. Human exposure to livestock – particularly through cohabitation – is associated with diarrheal disease and impaired child growth [3, 4]. Sixty-nine percent (20/29) of studies included in a systematic review and meta-analysis found a significant positive association between human exposures to domestic animals and diarrhea [3]. An observational analysis in rural Ethiopia determined that corralling poultry inside households overnight was associated with a 0.25 decrease (standard error = 0.118) in child height-for-age *Z*-scores [4]. Domestic animals are confirmed reservoirs of enteropathogens, including pathogenic *Escherichia coli, Campylobacter* spp., and *Salmonella* spp. [5–7]. Living in close proximity to animals, as is common in LMICs, is associated with increased presence of animal feces in households. [8–10]. Host-specific fecal markers from animals (dogs, birds, and ruminants) were detected in multiple household reservoirs (soil, hands, and stored drinking water) in rural and urban Bangladesh and rural India [10–12]. In rural Bangladesh, increased concentration of an animal-specific fecal marker, BacCow, was associated with increased prevalence of shiga toxin-producing *E. coli* on hands [11]. Living in close proximity to animals can therefore put humans at elevated risk of contact with animal feces, which can harbor enteropathogens.

Cohabitation between humans and domestic animals may also promote dissemination of antimicrobial resistance. Globally, by 2050, more than 10 million deaths due to antimicrobial-resistant infections are estimated to occur annually [13]. LMICs are predicted to bear the largest burden of these deaths, partially from poor regulation of antibiotic use in humans and animals, management of human and animal excreta, and access to safe drinking water, sanitation infrastructure, and hygiene [13]. Antimicrobial resistance arises through antibiotic use, accelerated by overuse and misuse of antibiotics (e.g., prophylaxis, incomplete courses, growth promotion) in humans and domestic animals, which contributes to the selection of intestinal bacterial strains that have acquired resistance [14]. Further, in microbial communities, such as the intestinal microbiota, bacteria can exchange antimicrobial resistance genes (ARGs) through horizontal gene transfer, allowing the exchange of ARGs between benign bacteria and pathogens, as well as between distinct microbial communities (humans, animals, environmental matrices) [15, 16]. The development of antimicrobial resistance in animal gut microbiomes in response to antibiotic use as prophylaxis or growth promotion and subsequent transmission to humans, particularly farm workers, has been demonstrated in intensive farming environments and high-income countries [17]. Particularly in LMICs, data on antimicrobial-resistant infections acquired in domestic environments, however, are limited. A study among six LMICs – Bangladesh, Bolivia, Ghana, India, Pakistan, and South Africa – found that more than half of community-acquired infections in children were resistant to antibiotics [18]. It is crucial to determine whether domestic animals raised through small-scale animal production and backyard farming are important reservoirs of antimicrobial resistance in LMICs to determine if community-level interventions – in addition to national-level regulation of antibiotic use in agriculture, veterinary medicine, and human medicine – could reduce the burden of antimicrobial-resistant infections.

While less than 30 % of urban Bangladeshi households own domestic animals, compared to the more than 70 % of rural households, a greater percentage of backyard and small-scale farms (having under 2.5 acres of land) exist in urban compared to rural communities [19, 20]. Although humans might be exposed to a greater number of animals in rural areas, limited space in urban areas makes it more difficult to separate animals and animal feces from humans living in the community. It is unclear how these differing levels of human exposure to animals might affect similarity in gut bacterial communities or intestinal resistomes (collections of all ARGs) between humans and animals in rural versus urban communities. Urban communities have been hypothesized as potential hotspots of antimicrobial resistance transmission [21]. Previous work has shown there is a higher proportion of antimicrobial-resistant infections among children from urban compared to rural communities [22–24]. As rural residents continue to migrate to urban areas, urban populations are expected to rise [25, 26]. Thus, it is increasingly important to characterize transmission pathways of antimicrobial resistance – and how they differ in rural versus urban settings – to design targeted and effective interventions.

While prior research in LMICs demonstrates the potential for gut bacteria and ARGs to be shared between humans and domestic animals, metagenomic studies examining resistomes have primarily relied on fragmented assemblies produced with short-read sequencing technologies, limiting their ability to link ARGs to specific bacterial species or locate ARGs on chromosomes or mobile genetic elements (e.g., plasmids, transposons). A study in rural Kenya found greater similarities in gut microbiota composition between children and cattle sharing a household compared to those from different households, though the results were not statistically significant [27]. A study in rural and peri-urban Peru found that 41 % of antibiotic resistance proteins shared by human, animal, and environmental samples were encoded in the same genetic context (neighboring core genes), indicating past horizontal gene transfer, though the short-read assemblies revealed that 11 % of unique antibiotic resistance proteins were encoded in multiple contexts [28]. Recent work, limited to the study of *E. coli* cultured from children and domestic animals in Ecuador, has shown that while clonal relationships exist, horizontal gene transfer via diverse plasmids may present a greater challenge in the dissemination of antimicrobial resistance [29, 30].

Technological advances in molecular methods, such as long-read sequencing, enable improved assemblies of complex metagenomes over those produced with short-read technologies, thus facilitating a deeper understanding of how antimicrobial resistance circulates in communities. In this study, we used a combination of 16S rRNA gene amplicon and long-read sequencing to characterize differential overlap in gut microbiomes and resistomes between humans and common domestic animals (chickens and goats) in rural versus urban Bangladesh. We also compared ARG abundance by fecal host type and urbanicity and identified potentially pathogenic bacterial species and plasmids carrying ARGs.

## Methods

### Study Sites

We collected human and animal fecal samples from one rural and one urban community in Bangladesh. The rural community, Mymensingh district, is located roughly 120 km north of Dhaka, the capital city of Bangladesh. Rural households are typically organized into compounds that share a common courtyard, sometimes including a water source and/or latrine. Rural households in Bangladesh are characterized by high levels of animal ownership, high levels of animals roaming indoors during the day and moderate levels sleeping inside at night, poor drinking water quality, limited household latrine ownership, and limited access to adequate handwashing stations [31–34]. The urban community, Tongi sub-district, is 15 km north of Dhaka. It is densely populated, with many families living in single-room homes and sharing a common stove, water source, and/or latrine with up to 15 other families. While less than 30 % of urban households own animals, a greater proportion of animal-owning households have less than 2.5 acres of land in urban compared to rural areas [9, 19, 20]. Similar to rural areas, low-income households in urban Bangladesh are characterized by poor drinking quality and insufficient sanitation facilities [35, 36]. According to recent studies, reported household antibiotic use among children under 5 years is fairly similar in rural (39.1 %) and urban (44.2 %) communities in Bangladesh [36, 37].

### Sample Collection

Between June and August 2015, 50 individual chicken and 50 individual goat fecal samples were identified by observation in public and domestic areas in Bangladesh, half of which were from Mymensingh (rural) and half from Tongi (urban). Using sterile scoops, fresh feces were scooped from the center and top of feces piles to avoid soil contamination. Samples were stored on ice and transported to a field laboratory (Mymensingh) or International Centre for Diarrhoeal Disease Research, Bangladesh (icddr,b; Tongi) within six hours. Trained icddr,b staff carried out the animal fecal sample collection.

Twenty-five human stool samples were collected from adult (>18 years) residents of Tongi between April and October 2013 [38]. The samples were transported in Cary-Blair media to icddr,b and frozen at -80 °C. Between June and September 2015, 20 human stool samples were collected from adult (>18 years) residents of Mymensingh [39]. The samples were transported to the field laboratory and frozen at -80 °C before being shipped to icddr,b on dry ice.

Animal fecal samples were immediately processed after arrival to the laboratories, and human stool samples were thawed from -80 °C prior to further processing. The samples were combined into fecal composites for each fecal host (chicken, goat, human) by measuring 0.2 g of five individual samples from corresponding locations into 50-mL centrifuge tubes using ethanol and flame sterilized spatulas. After mixing the samples, 1-g composites were transferred to 1.5-mL microcentrifuge tubes. 1 mL of RNALater™ was mixed into each composite and left to sit for five minutes. Five composites were made for each fecal host in each community (rural, urban), with the exception of only four composites for rural human stool. Chicken fecal composites were treated in a hot water bath (100 °C) for 20 minutes in accordance with our United States Department of Agriculture (USDA) import permit (128693). All composites were stored at -80 °C before shipment at room temperature to Stanford University, where samples were stored at - 80 °C upon arrival. Aliquots for the 16S rRNA gene analysis were stored at -80 °C (stored 2 years and 9 months before processing) at Stanford University. After storage at Stanford University, aliquots for the long-read sequencing analysis were shipped on dry ice to Tufts University and stored at -80 °C (stored an additional 4 months before processing).

### DNA Extraction

For 16S rRNA gene analysis, DNA was extracted from 0.25 g of each fecal composite using the MoBio PowerSoil® extraction kit (Carlsbad, CA) according to the manufacturer’s instructions. We extracted one blank sample per day of DNA extractions (three extraction controls total) to control for contamination during the extraction process. *E. coli* K-12 (ATCC 10798) was used as a positive control. The culture was grown on tryptic soy agar and a single colony was suspended in nuclease-free water (Thermo Fisher Scientific, Waltham, MA). The suspension was centrifuged at 10,000 x g for 5 minutes, and the pellet was resuspended in nuclease-free water, heated to 100 °C for 10 minutes, cooled at 20 °C for 10 minutes, and centrifuged at 14,000 x g for 5 minutes. The supernatant was used as template in polymerase chain reactions (PCRs).

Due to budget constraints, only four of the five fecal composites from each fecal host were processed with long-read sequencing. For long-read sequencing, DNA was extracted from 0.25 g of each fecal composite using the DNeasy® PowerSoil® Pro extraction kit (QIAGEN, Germantown, MD). We followed the manufacturer’s instructions, with the exceptions of using an alternate lysis method (heating samples at 70 °C for 10 minutes followed by vortexing at full speed for 10 minutes, adapted from Earth Microbiome Project’s DNA extraction protocol) and eluting in Tris-EDTA buffer (Integrated DNA Technologies, Inc., Coralville, IA) to prevent degradation [40]. We extracted one blank sample per day of DNA extractions (three extraction controls total). DNA purity was quantified using a NanoDrop (Thermo Fisher Scientific, Waltham, MA). For extraction controls, nucleic acid concentrations were measured using a Qubit™ 4.0 fluorometer (1X dsDNA HS assay kit; Thermo Fisher Scientific, Waltham, MA). Nucleic acid concentration and fragment size distributions for fecal composite extracts were assessed using an Agilent 2100 Bioanalyzer at the Tufts University Core Facility (Boston, MA). Quality control parameters are presented in Table S1.

### 16S rRNA Gene Amplicon Generation and Sequencing

We used a previously described 16S rRNA gene sequencing Illumina amplicon protocol with 515f and 805r primers (GTGYCAGCMGCCGCGGTAA and GGACTACNVGGGTWTCTAAT, respectively) that targeted the V4 region of the 16S rRNA gene [41–46]. Golay barcodes were embedded in each forward primer (Table S2). Previously described PCR protocols were used to generate 16S rRNA gene amplicons from all samples, including the negative and positive controls: 25 μL PCRs consisted of 2x QIAGEN HotStarTaq® Plus mastermix (Germantown, MD), 10 μM of forward primer, 10 μM of reverse primer, and 1 μL of template DNA. PCRs were run in triplicate using the following thermocycler conditions: 94 °C for 3 minutes; then 35 cycles of 94 °C for 45 seconds, 50 °C for 60 seconds, 72 °C for 90 seconds; followed by 72 °C for 10 minutes and a hold at 4 °C [41–45]. A no template control reaction was included in every PCR run. After PCR, we visualized product size using electrophoresis on 1.5 % agarose gels containing ethidium bromide. All fecal composites and positive controls showed a band of appropriate size (∼390 bp). The PCR triplicates each received unique barcodes during PCR (i.e., each sample was sequenced in triplicate). We did not observe any bands in the lanes of the gel containing the negative controls, including extraction and no template controls.

A Qubit™ 2.0 fluorometer (dsDNA HS assay kit; Thermo Fisher Scientific, Waltham, MA) was used to quantify the nucleic acid concentration in PCR products. Based on those concentrations, amplicons from the samples and the positive control were multiplexed and pooled in equimolar proportions (10 µM) for sequencing. Additionally, we multiplexed and pooled extraction and no template controls (negative controls) for sequencing. 10 µL of each of the negative control pools were added to the total pool before purification. The total pool was purified using the MoBio UltraClean® PCR clean-up kit (Carlsbad, CA) before sequencing. Sequencing was performed using an Illumina MiSeq at the Stanford Functional Genomics Center (Stanford, CA) [46]. We used 250-bp paired-end reads (2×250) and spiked the total pool with 10 % phiX before sequencing. We sent index primers (AATGATACGGCGACCACCGAGATCTACACGCT), read 1 primers (TATGGTAATTGTGTGYCAGCMGCCGCGGTAA), and read 2 primers (AGTCAGCCCAGCCGGACTACNVGGGTWTCTAAT) with the total pool [41–45].

### Long-Read Sequencing

Long-read sequences were obtained from DNA extracts using Oxford Nanopore Technologies’ (ONT’s) MinION™. Library preparation was conducted using the SQK-LSK109 ligation sequencing kit (ONT, Oxford, United Kingdom) according to the manufacturer’s instructions. When preparing libraries for goat and human samples, the L Fragment Buffer (LFB) was used to enrich for DNA fragments of 3 kbp or longer. Since DNA concentrations were low in chicken fecal composite extracts (Table S1), however, the S Fragment Buffer (SFB) was used to retain DNA fragments shorter than 3 kbp. Two fecal composite libraries were multiplexed per flow cell (FLO-MIN106; ONT, Oxford United Kingdom) in equimolar proportions (except for chicken samples, for which all DNA was retained to account for the lower DNA concentrations) using the EXP-NBD103 native barcoding kit (ONT, Oxford, United Kingdom).

### Data Processing and Analysis

#### 16S rRNA Gene Amplicon Analysis

The DADA2 pipeline was used to process forward and reverse reads [47]. Forward reads were truncated to 173 nucleotides and reverse were truncated to 162 nucleotides, after which the quality score dropped. Paired-end reads were merged to yield 250 bp sequences, and chimeras were removed. Taxonomy was assigned in DADA2 using a Naïve Bayes classifier that was trained on the Silva v132 database [48, 49]. Species-level identification was based on 100% identity between the reference database and amplicon sequence variants (ASVs) [50].

Data were analyzed using *phyloseq* (version 1.26.1) in R (v3.5.0) [51]. All ASVs associated with eukaryotic organelles – chloroplasts and mitochondria – were removed. ASVs were normalized using the inverse hyperbolic sine transformation [52]. ASVs identified in the 3 replicates were merged by summing, and the merged data were used for all subsequent analyses. To determine the similarity of bacterial communities between humans and animals, we calculated pairwise Bray-Curtis dissimilarity indices between humans and goats and between humans and chickens in rural and urban areas. A Wilcoxon rank sum test (*p*<0.05) was used to determine if the differences in Bray-Curtis indices were statistically significant in urban versus rural areas. In a sensitivity analysis, ASVs classified as the 140 taxa present in the no template control (Figure S1) were removed from all samples and Bray-Curtis dissimilarities were recalculated. Variables contributing to differences between communities, such as fecal host and location, were identified by PERMANOVA with Bray-Curtis dissimilarity using *vegan* (v2.5.6) in R (v3.5.0) [53].

#### Long-Read Sequencing Analysis

Long-read sequences were basecalled, demultiplexed, and trimmed using ONT’s toolkit, Guppy (v3.3.0). To identify ARGs, reads were aligned to the ResFinder database (duplicates removed) using Minimap2 (v2.15) [54, 55]. Only primary ARG alignments with ≥90 % similarity and ≥100 bp alignment lengths were retained for the analysis. To quantify data classified as bacteria, raw reads were aligned to the National Center for Biotechnology Information’s (NCBI’s) RefSeq database (plasmid sequences removed) using a least common ancestor approach in Centrifuge (v1.0.4) [56]. ARG counts were normalized by gigabase pairs (Gbp) of data classified as bacteria. Wilcoxon matched-pairs signed-rank tests (*p*<0.05) were used to test for statistical differences in the normalized abundance of ARGs, paired by drug class, in urban and rural samples by fecal host. Wilcoxon rank sum tests (*p*<0.05) were used to test for differences in the normalized abundance of ARGs in human, chicken, and goat samples by urbanicity. To test the hypothesis that human and animal resistomes were more similar in rural versus urban communities, Bray-Curtis dissimilarities were calculated for all pairwise comparisons, and a Wilcoxon rank sum test (*p*<0.05) was used to determine statistical significance. We identified overlapping distinct resistance gene alleles between humans and each animal within urban and within rural communities by generating ARG overlap matrices and using two proportion *Z*-tests (*p*<0.05) to determine statistical significance in rural versus urban areas.

To identify potential pathogens carrying ARGs, reads were assembled using Flye (v2.6) (with the meta option) and polished using ONT’s tool for sequence correction, medaka (v0.11.0) [57]. Polished contigs were aligned to the modified RefSeq database using Centrifuge and modified ResFinder database using Minimap2 (v2.15) [54–56]. We focused our analysis on the 12 bacterial pathogens identified as antimicrobial-resistant threats in the Centers for Disease Control and Prevention (CDC) 2019 Antibiotic Resistance Threats in the United States report: *A. baumanni, C. coli, C. jejuni, C. difficile, Enterococcus faecium, E. coli, K. pneumoniae, N. gonorrhoeae, S. enterica, S. dysenteriae, S. aureus*, and *S. pneumoniae* [58]. The reliability of pathogen classifications at the sub-species and strain level carrying resistance genes was assessed by aligning potential pathogen sequences against their respective classifications in NCBI’s RefSeq database using BLASTN; an *E*-value that rounded to 0 (BLASTN uses double-precision floating-point format, therefore the smallest *E*-value reported is about 1e-179) and percent identity ≥98 % were the thresholds used to confirm whether a pathogen classification was reliable [59]. Plasmids were identified from the contigs using the MOB-suite tool to determine whether identified ARGs were chromosome-or plasmid-associated [60]. Long-read data from the same type of fecal host and location were pooled together for presentation of the results. Statistical analyses and data visualization were conducted using Stata SE (v14.2) and R (v3.6.3).

## Results

### Similarity in Gut Microbiomes

We obtained a total of 11.7 million reads from the 16S rRNA gene amplicon sequencing of DNA extracts from chicken, goat, and human fecal composites. The average sequencing depth per sample was 97,000 reads in chickens, 134,000 in goats, 144,000 in humans, 72,000 in the positive control (*E. coli* K-12 (ATCC 10798)), 180 in the extraction blank, and 6,600 in the PCR no template control (nuclease-free water in place of template DNA). On average, 79 % of the input reads remained after quality filtering, merging forward and reverse reads, and chimera removal. Sixty-six percent and 8 % of reads were classified at the genus and species level, respectively. In total, 1,528 ASVs were present in chicken fecal composites, 3,270 in goat composites, and 655 in human composites. The relative abundance of the 30 most abundant families in all sample types are presented in Figure 1. There were 140 ASVs identified in the no template control (Figure S1) indicating potential contamination during sample processing. We performed a sensitivity analysis in which the ASVs present in the no template control were removed from all samples before proceeding with data analysis (Figure S2). Due to the absence of standardized methods to account for negative controls and our finding that the primary study conclusions remained unchanged in the sensitivity analysis, we did not adjust fecal sample ASV counts in our primary analysis.

**Figure 1.**
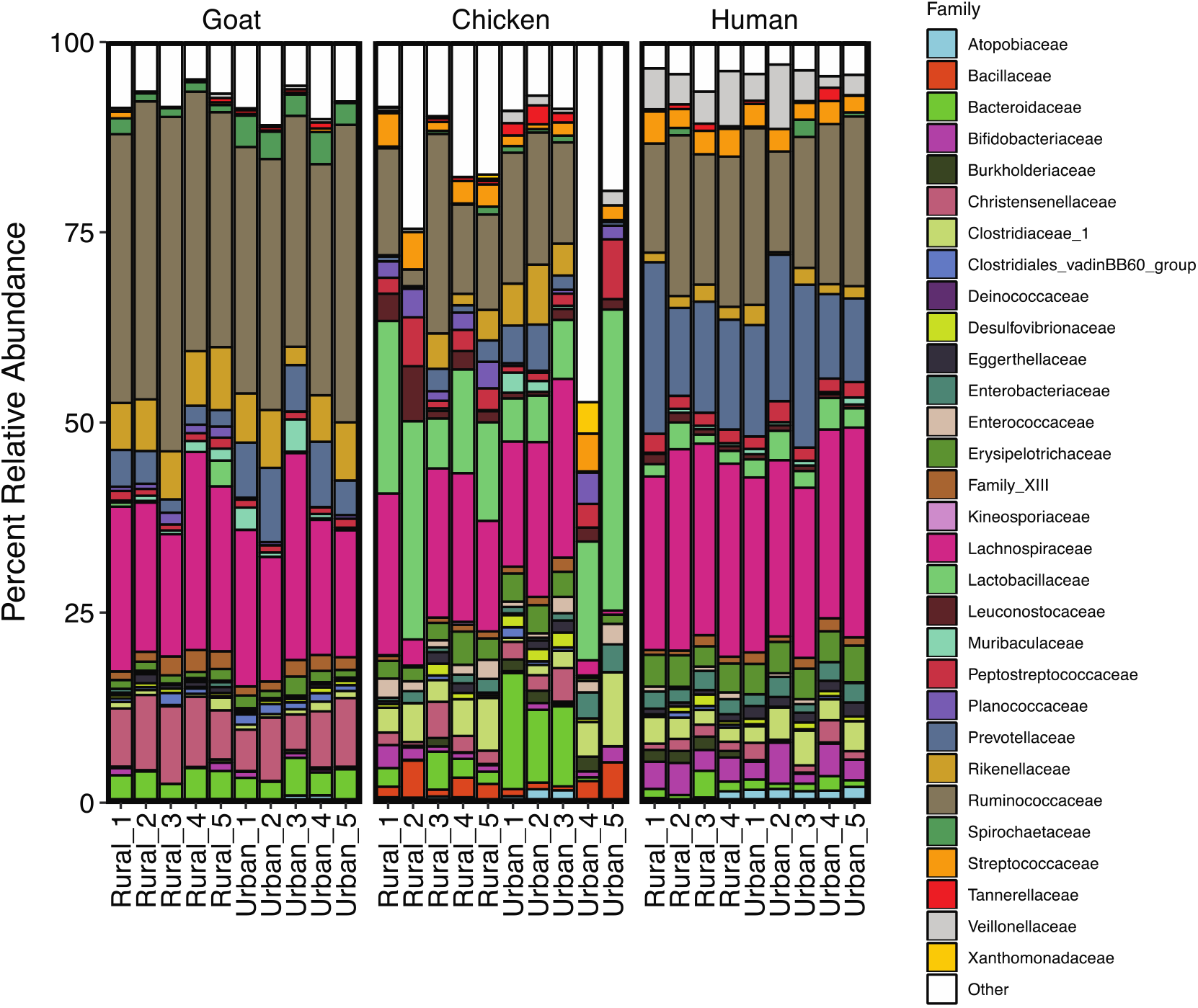
Relative abundance of 30 most abundant families in all samples from 16S rRNA gene analysis. All other taxa are grouped into “Other”.

The bacterial community compositions of human, chicken, and goat feces were significantly different by fecal host (PERMANOVA *R*^*2*^=0.50, *p*=0.001) (Figure 2). Urbanicity was an important determinant for the gut bacterial community composition in humans and in goats, but not in chickens. Specifically, within each fecal host, rural and urban bacterial community compositions were significantly different in goats (*R*^*2*^=0.24, *p*=0.01) and humans (*R*^*2*^=0.20, *p*=0.02) but not in chickens (*R*^*2*^=0.14, *p*=0.19). While microbiomes of humans and animals were distinct, human microbiomes were more similar to those in chickens than in goats. The similarity of human and chicken microbiomes was greater in rural versus urban fecal samples (Bray-Curtis dissimilarity index median [IQR] rural=0.92 [0.03], urban=0.94 [0.01]; Wilcoxon rank sum *p*<0.001) (Figure 2). The similarity between human and goat fecal bacterial community compositions, however, did not differ in rural versus urban areas (rural=0.97 [0.04], urban=0.95 [0.03]; Wilcoxon rank sum *p*=0.30).

**Figure 2.**
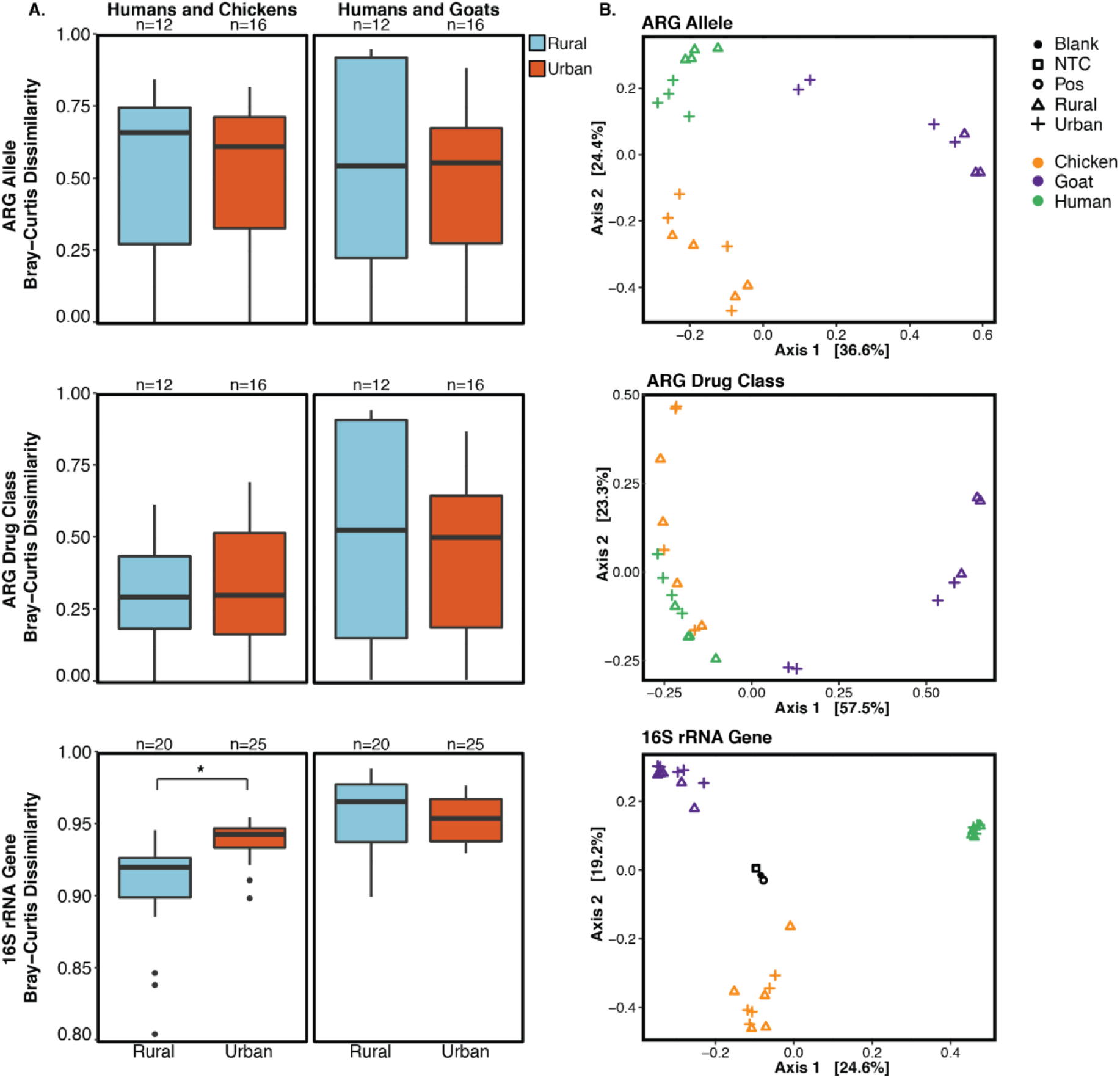
Bray-Curtis dissimilarity between resistomes (top and middle; long-read sequencing) and bacterial community compositions (bottom; 16S rRNA gene analysis) in humans, chickens, and goats. **A**. Box-and-whisker plots illustrating Bray-Curtis dissimilarity between humans and chickens (left) and humans and goats (right). The line through each box denotes the median Bray-Curtis dissimilarity index, the lower and upper box boundaries denote the first and third quartiles, respectively, and the whiskers extend to the minimum and maximum indices up to 1.5 times the interquartile range, with outliers plotted as dots. A Bray-Curtis dissimilarity index equal to 0.00 indicates identical community compositions and closer to 1.00 indicates more dissimilarity between community compositions. **B**. Principal coordinate analyses (PCoAs) of Bray-Curtis dissimilarity. Color indicates fecal host and symbol indicates rural, urban, or control.

### Similarity and Overlap in Resistomes

The long-read sequencing of DNA extracts from the fecal composites yielded, on average, 8.8 million reads per sample type (fecal host by urbanicity) with an average read length of 1,872 bp (standard deviation: 2,546) (Table S3). We obtained a total of 99 Gbp of data, 71 % of which was classified as bacteria. Assembly yielded an N50 of 50 kbp, on average, and a median contig length of 25 kbp.

We identified a total of 44,077 ARG hits and 300 distinct ARG alleles. Tetracycline, beta-lactam, and macrolide, lincosamide, and streptogramin B (MLS) resistance genes dominated human and animal resistomes (Figure 3). ARGs were widespread, especially in chicken and human feces. The abundance of ARGs (normalized by Gbp of data classified as bacteria for each fecal host) was greater in chickens (Wilcoxon rank sum test, rural: *p*=0.034, urban: *p*=0.021) and in humans (Wilcoxon rank sum test, rural: *p*=0.034, urban: *p*=0.021) compared to goats (Table S4). The abundance of ARGs was higher in urban versus rural animals, but not significantly different by urbanicity for human guts. Specifically, within each fecal host, ARG abundance (paired by drug class) was greater in urban compared to rural samples, though results were only statistically significant for chickens and goats (Wilcoxon signed-rank test, chickens: *p*=0.046, goats: *p*=0.017, humans: *p*=0.29; Figure 3, Table S5). In rural versus urban areas, the similarity between human and chicken resistomes at the allele level (rural=0.66 [0.47], urban=0.61 [0.39]; Wilcoxon rank sum *p*=0.28) and grouped by drug class (rural=0.30 [0.25], *p*=0.58) did not differ (Figure 2). Similarities between human and goat resistomes at the allele level (rural=0.55 [0.70], urban=0.56 [0.40]; *p*=0.17) and grouped by drug class (rural=0.53 [0.77], urban=0.50 [0.46]; *p*=0.12) also did not differ (Figure 2).

**Figure 3.**
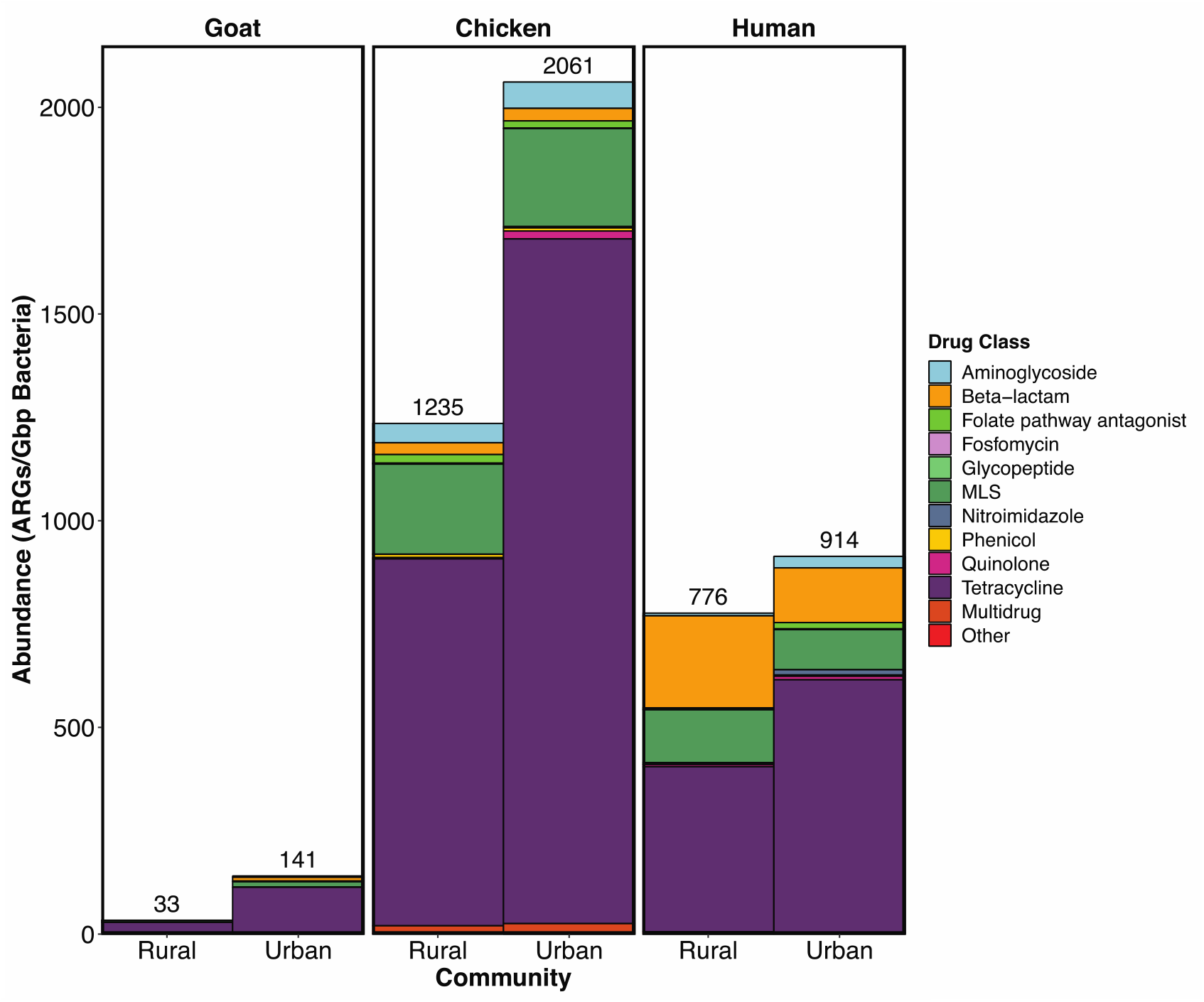
Abundance of antimicrobial resistance genes (ARGs) (identified in long-read sequencing data) normalized by Gbp of data classified as bacteria for each fecal host by urbanicity. MLS represents macrolide, lincosamide, and streptogramin B resistance genes.

Overlap of distinct ARG alleles was greater between humans and goats (*Z*-test *p*=0.017) in the urban compared to rural community, though not significantly different between humans and chickens (*Z*-test *p*=0.212) in urban versus rural samples (Figure 4). In rural and urban communities, the overlap of ARGs in humans and chickens was greater than that in humans and goats (*Z*-test *p*<0.001). Among genes overlapping between urban chickens and urban humans, three extended-spectrum beta-lactamase (ESBL) genes, *bla*_*TEM*_*-126, bla*_*TEM*_*-132*, and *bla*_*TEM*_*-207*, and three beta-lactamase inhibitor (BLI)-resistant genes, *bla*_*TEM*_*-32, bla*_*TEM*_*-76*, and *bla*_*TEM*_*-122*, were identified. In addition, *blaTEM-76* overlapped in rural chickens and rural humans. While not overlapping between fecal hosts living in the same community, additional ESBL genes, *bla*_*SHV*_*-12* and *bla*_*TEM*_*-147*, were detected in chicken and human hosts. Notably, *floR*, a gene conferring resistance to florfenicol, was detected in rural humans and urban chickens.

**Figure 4.**
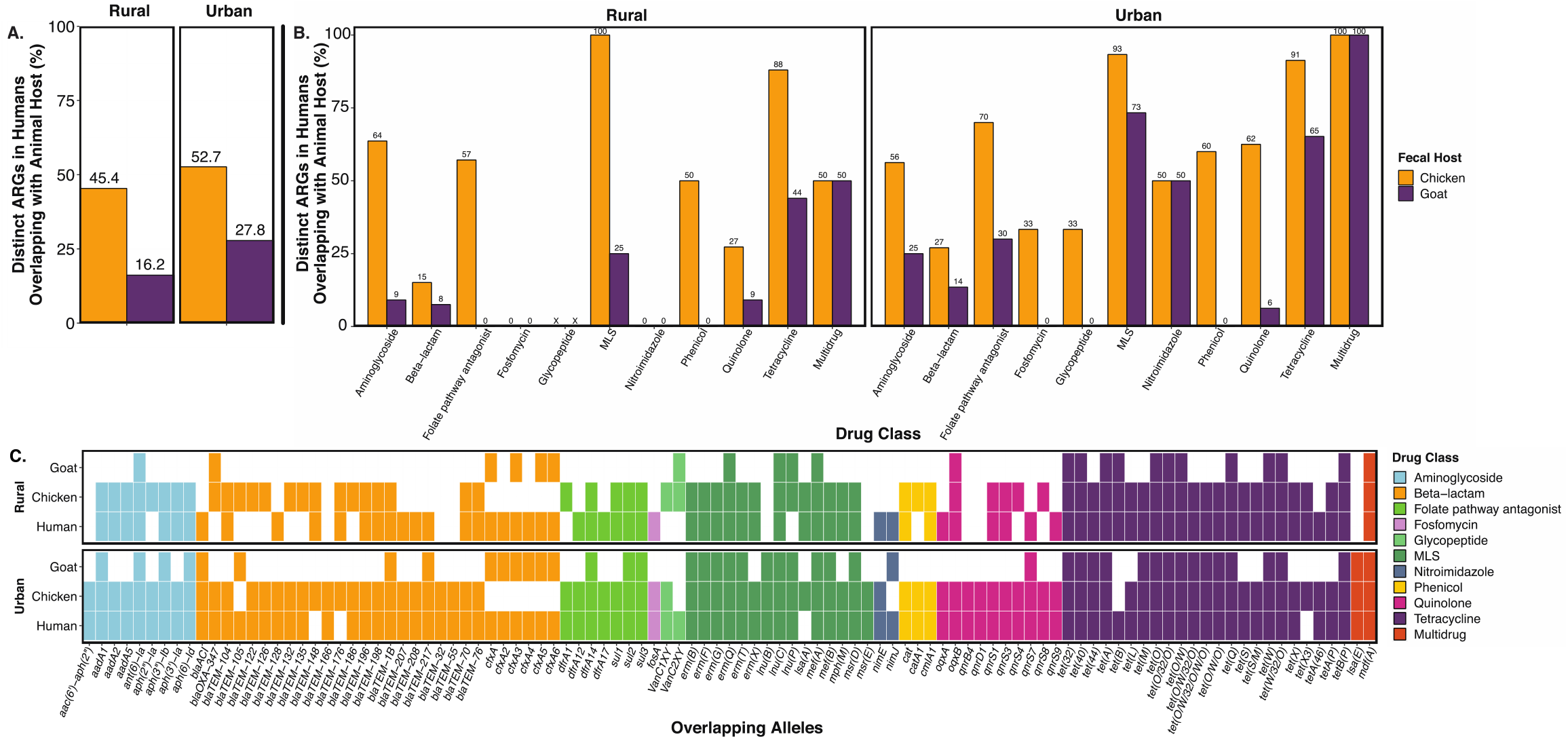
Distinct antimicrobial resistance gene (ARG) alleles overlapping between humans and each animal host. **A**. Percentage of distinct ARGs identified (using long-read sequencing) in humans also found in each animal host. **B**. Percentage of distinct ARGs, by drug class, identified (using long-read sequencing) in humans also found in each animal host. An ‘X’ indicates ARGs from that drug class were not detected in humans from that community. **C**. ARG alleles overlapping between at least two fecal hosts living in the same community.

We identified plasmids carrying resistance in the urban chicken, rural human, and urban human samples (Table 1). One plasmid cluster was shared by humans and chickens, but it could not be typed. A total of seven unique plasmid types (ColRNAI, Inc18, IncFIB, IncR, IncFII, IncFIIA, IncQ1) were present in the rural and urban human samples, and none of the plasmids identified in urban chicken samples could be typed. *Bla*_TEM_ genes conferring resistance to beta-lactams were carried by IncF-type and ColRNAI plasmids in human stool.

**Table 1.**
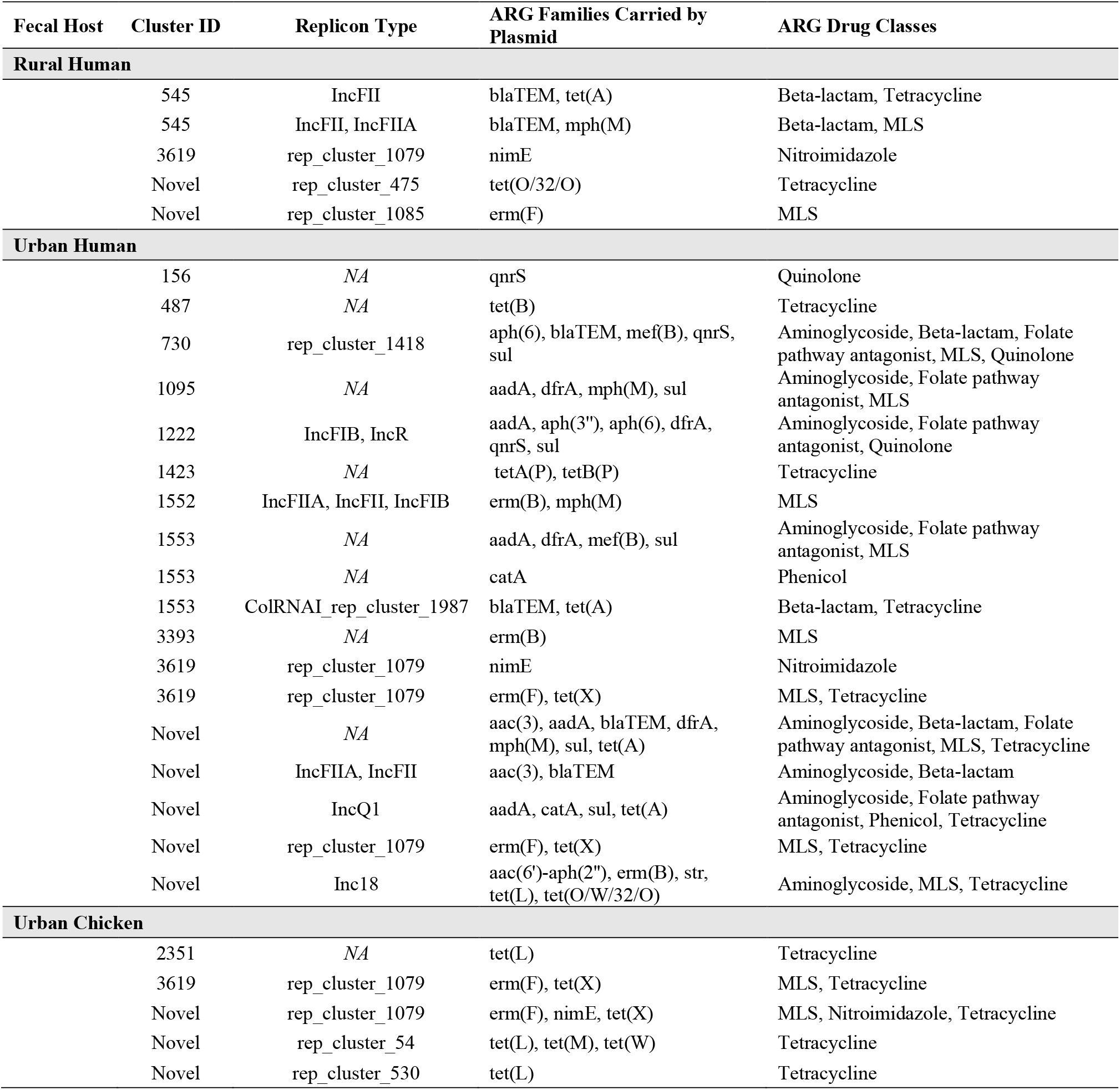
Plasmids carrying antimicrobial resistance genes. Some fecal hosts (rural chicken, rural goat, urban goat) are not shown, because no plasmids carrying antimicrobial resistance genes were detected in their respective samples.

Of the contigs assembled from the long reads, 87.5 % (99,823/114,093) were classified as bacteria. Eighty-nine percent (89,228/99,823) of the bacterial contigs were classified to the strain, subspecies, or species level, of which 2.37 % (2,112/89,228) were classified as potentially pathogenic species highlighted in the CDC 2019 Antibiotic Resistance Threats report [58]. We determined that several important pathogens were likely present and carried antimicrobial resistance, particularly in the urban community (Figure 5). In total, 28 distinct resistance genes, representing 10 drug classes (aminoglycoside; beta-lactam; fluoroquinolone; folate pathway antagonist; fosfomycin; macrolide, lincosamide, and streptogramin B; phenicol; quinolone; rifampicin; and tetracycline), were carried by potential pathogens. No resistance genes were carried by potential pathogens in rural chickens or rural goats. Of the six bacteria species carrying resistance in humans, two were also present in urban animal hosts. *C. difficile* organisms carrying tetracycline (*tet(W), tet(O), tet(40)*) and macrolide (*mef(A)*) resistance were present in humans and urban goats, and *C. difficile* organisms carrying aminoglycoside (*ant(6)-Ia*) resistance were present in urban humans and urban chickens. Multidrug resistance (*mdf(A)*) carried by *E. coli* was prevalent in humans, urban chickens, and urban goats. *E. coli* carrying beta-lactam resistance (*bla*_TEM_) were identified in urban chickens and urban humans. Only three contigs carrying resistance genes were classified as potential pathogens to the strain or subspecies level. *E. coli PCN033* carrying multidrug resistance (*mdf(A)*) was identified in a rural human sample, and *C. difficile M120* carrying macrolide (*mef(A)*) and tetracycline (*tet(44)*) resistance were present in an urban goat and urban human sample, respectively. Though the potential pathogen classifications were confirmed through BLASTN with low *E*-values and high percent identity (Table S6), we noted off-target hits with similar *E*-values and percent identities for the *E. coli PCN033* and *C. difficile M120* classifications.

**Figure 5.**
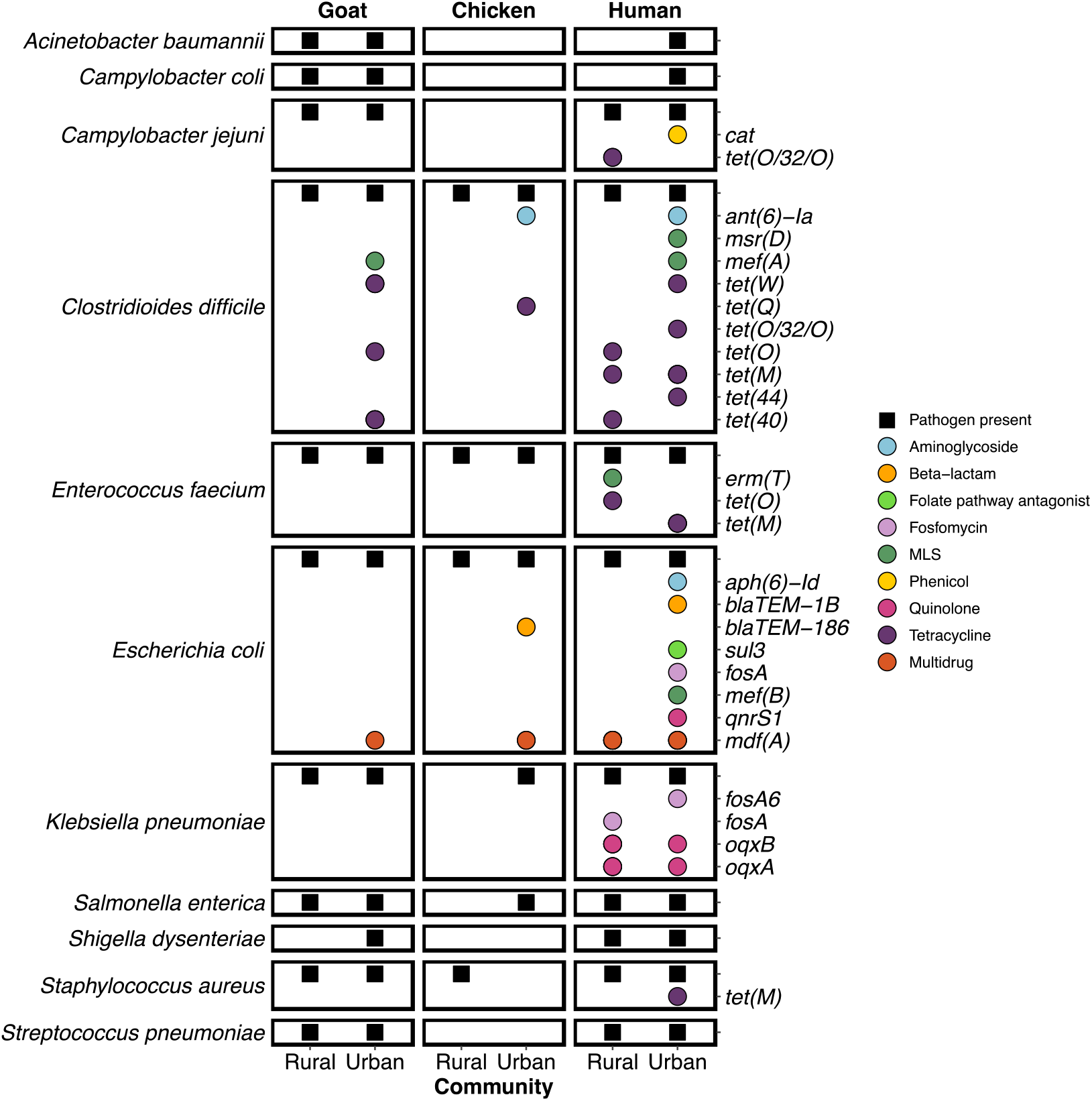
Prevalence of potentially pathogenic bacteria species and those carrying antimicrobial resistance genes (ARGs) identified in contigs assembled from long reads. Black squares indicate at least one contig for the respective fecal host was classified as the bacteria species on the left. Colored dots indicate ARGs (by drug class) identified on contigs classified as the bacteria species listed on the left. Bacteria species represent those identified in the CDC’s 2019 Antibiotic Resistance Threats in the United States report (31). MLS represents macrolide, lincosamide, and streptogramin B resistance genes.

## Discussion

We found greater similarity in bacterial communities between humans and chickens in rural versus urban Bangladesh, which could be explained by a higher prevalence of animal ownership in the rural site. While more than 70 % of households own livestock in rural Bangladesh, less than 30 % own animals in urban areas [19]. As our study did not specifically collect human stool from animal-owning households, it is possible human participants from the urban community had limited exposure to animals. Additionally, human microbiomes are generally more diverse in urban compared to rural areas [61]. Findings from previous studies have shown that gut microbiota are influenced by geography and diet [28, 61–64]. People living in urban Bangladesh tend to have diverse diets consisting of most food groups (i.e., starches, pulses, fish, eggs, meat, vegetables, fruit, milks, oil, spices, sweets, beverages), while those in rural areas are more likely to have limited diets [65]. Geography and diet could also explain why urbanicity was a determinant of bacterial communities in humans and in goats. Free-roaming animals are common in rural and urban Bangladesh, so it is possible the differences in bacterial communities between rural and urban goats could be attributed to grazing on differing food sources in rural and urban communities [8, 66, 67].

Our findings highlight domestic chickens as important reservoirs of antimicrobial resistance. Similarly, a comparison of resistomes in chickens, pigs, and humans in China found ARG abundance in chicken feces to be the highest [68]. Another study found that publicly available human stool bacterial genomes shared a greater number of mobile ARGs with chicken gut microbiomes than with pig and cattle microbiomes [69]. The high abundance of ARGs in chicken feces and resistome sharing between chickens and humans is consistent with existing literature, much of which has focused on occupational exposures in commercial settings. For example, recent work in Nigeria identified occupational exposure to chickens as a risk factor for multidrug-resistant *E. coli* among poultry workers and identified similar resistance patterns and identical plasmid replicons in *E. coli* recovered from chickens and poultry workers [70–72].

While previous work in Ecuador found that higher levels of antimicrobial-resistant bacteria were isolated from commercially raised poultry than backyard poultry, our work uniquely demonstrates the important role of poultry in antimicrobial resistance circulation in urban communities [73]. Interestingly, we detected *floR* in not only urban chickens, but rural humans as well. Florfenicol is generally used in veterinary medicine, in aquaculture, and as growth promotion for livestock, but is not approved for human use [74]. As in a 2021 study that found florfenicol resistance levels to be higher in chicken vendors than the general population, detection of *floR* in human stool may indicate antimicrobial resistance originating from animal sources [75].

Tetracycline resistance genes dominated the resistomes of humans, chickens, and goats. The tetracycline class includes inexpensive antibiotics that have been widely used as prophylaxis and to treat a variety of gram-positive and gram-negative bacterial infections in humans and domestic animals; tetracyclines have also been used as growth promoters in animal husbandry [76]. Our results mirror previous work that found tetracycline resistance genes to be among the most abundant ARGs in human and animal feces [68, 77–79]. In layer farms in Bangladesh, tetracycline was one of the most prevalent antibiotic residues, in addition to ciprofloxacin and enrofloxacin (fluoroquinolones), in the eggs, tissue, and internal organs of chickens [80, 81]. Genes conferring resistance to MLSs were also abundant in our study, particularly in humans and chickens. Similar to tetracyclines, MLSs have a long history of prophylactic and treatment use in human and veterinary medicine [82]. Additionally, beta-lactam resistance genes, including ESBL and BLI-resistant genes, were prevalent in our study samples, especially in chickens and humans. Beta-lactamases, particularly cephalosporins, were one of the most widely prescribed antibiotics in multiple studies surveying pharmacies, outpatients, and hospitals in rural and urban Bangladesh [83–86]. ESBL-producing *Enteriobacteriaceae* infections are a global public health concern and have limited treatment options, often requiring last-resort treatment with carbapenems [87]. In addition to other gram-negative bacterial infections, ESBL-producing *Enteriobacteriaceae* infections are often treated by co-administering beta-lactams and beta-lactamase inhibitors [88]. Further, beta-lactamase enzymes are widely plasmid-mediated, allowing for ease of horizontal gene transfer between bacterial hosts and between humans, animals, and the environment [16, 89, 15, 90].

We detected *bla*_TEM_ genes (beta-lactam resistance) located on IncF plasmids in rural and urban human stool and on a ColRNAI plasmid, a plasmid group known to carry beta-lactam resistance in *Enterobacteriaceae*, in urban human stool [89, 91–94]. IncF plasmids are particularly concerning in the spread of antimicrobial resistance, as they often carry multiple replicons, or regions of the genome where replication is initiated, allowing for rapid evolution and great adaptability to promote acquisition by various bacterial host species [93]. Although there is a recognized importance of mobile genetic elements (e.g., plasmids) in horizontal gene transfer and the spread of antimicrobial resistance, we detected few plasmids in our long-read sequencing data. Accurate identification of plasmids using the MOB-suite tool, as well as other plasmid detection tools, relies on high quality genome assemblies as input [60]. Despite advances in the accuracy of long-read sequencing technology, sequencing errors and insufficient coverage make plasmid detection difficult in complex metagenomes using long reads alone. Deeper sequencing and hybrid assemblies of short- and long-read data may improve the ability to characterize the role of plasmids in human and animal resistomes. Further, MOB-suite uses a database-based approach, and performance is reduced when classifying novel plasmids not available in NCBI’s RefSeq and GenBank databases [60]. Due to overrepresentation of plasmids from human clinical isolates from high-income countries in databases, it is possible that we were limited in our ability to detect plasmids in chickens and goats or specific to Bangladesh using a database approach.

We leveraged long sequence reads to identify bacterial hosts of ARGs. *C. difficile* carried a wide range of ARGs across various drug classes (aminoglycoside, MLS, and tetracycline) in humans and animals. *C. difficile* can colonize the gut in spore form without causing illness, particularly in infants; asymptomatic colonization in adults is rare, but possible [95–97]. Historically, *C. difficile* infections have been known to arise after taking antibiotics and account for the highest morbidity and mortality of hospital-acquired infections in high-income countries, though the burden of *C. difficile* infections in LMICs is poorly understood [98–101]. There is, however, increasing recognition of *C. difficile* as a community-acquired infection, in addition to hospital-acquired infection, and potential zoonosis [102, 103]. We also identified *E. coli* carrying *bla*_TEM_, a gene family containing alleles conferring resistance to beta-lactams, in urban chickens and urban humans. Previous work has documented poultry as reservoirs of ESBL-producing *Enterobacteriaceae* [104, 105]. Our results therefore stress the importance of understanding and targeting transmission pathways of ESBL-producing *Enterobacteriaceae* in curbing antimicrobial resistance, particularly in urban areas where humans and animals live in close proximity and share environments.

This study has some limitations. We leveraged human stool collection from previous studies, and the human and animal samples were not temporally matched in the urban community. We are unable to draw conclusions about whether zoonotic transmission occurred (or its directionality) in our study communities. In addition, few barcodes were detected in the long-read sequence data for one of the rural goat samples, so we were not able to detect any ARGs, plasmids, or potential pathogens in that sample; however, as all long-read sequencing analyses (excluding the Bray-Curtis dissimilarity analysis) relied on pooled data from each fecal host in a particular setting, we do not anticipate the missing sample should influence the interpretation of our results. When comparing potential pathogens detected in the chicken feces versus other fecal hosts, it should be noted that we obtained less and more fragmented long sequence reads from chicken DNA extracts compared to human and goat DNA, potentially due to the heterogeneity, heat treatment (required by our USDA import permit), or high uric acid concentration of chicken fecal matter.

## Conclusions

This community-level study demonstrates similarities in resistance profiles, as well as reveals potential pathogens carrying resistance, in humans and animals living in the same community in rural and urban Bangladesh. Our results contribute to a gap in knowledge regarding the role of small-scale animal production in antimicrobial resistance dissemination within resource-constrained communities. While domestic goats and chickens harbored potentially pathogenic hosts of ARGs, our results suggest that compared to goats, backyard poultry might be an especially important reservoir for ARGs that are also present in the human gut. In addition, our findings provide some evidence that ARGs are more widespread in urban compared to rural communities. Substantial sharing of ARGs in humans and animals in both rural and urban communities in this study highlights the importance of developing interventions and antimicrobial stewardship strategies that account for both human and animal reservoirs.

## Data Availability

Metagenomic sequence reads and 16S rRNA amplicon sequences are available in the sequence read archive (SRA) under accession numbers SRR13059261- SRR13059374. Human sequences were removed from all samples. Relevant analysis scripts are available in GitHub (https://github.com/jennaswa/arg_bd).

## List of Abbreviations

LMIC: low- and middle-income country
ARG: antimicrobial resistance gene
USDA: United States Department of Agriculture
PCR: polymerase chain reaction
ONT: Oxford Nanopore Technologies
ASV: amplicon sequence variant
NCBI: National Center for Biotechnology Information
CDC: Centers for Disease Control and Prevention
MLS: macrolide, lincosamide, and streptogramin B
ESBL: extended-spectrum beta-lactamase
BLI: beta-lactamase inhibitor

## Declarations

### Ethics Approval and Consent to Participate

Written informed consent was obtained from all study participants. The studies were approved by the icddr,b Ethical Review Committee (PR-11063, PR-13004), Johns Hopkins University Institutional Review Board (IRB) (00004795), University of California, Berkeley Committee for the Protection of Human Subjects (2011-09-3652), and Stanford University IRB (25863).

### Consent for Publication

Not applicable.

### Availability of Data and Materials

Metagenomic sequence reads and 16S rRNA amplicon sequences are available in the sequence read archive (SRA) under accession numbers SRR13059261-SRR13059374. Human sequences were removed from all samples. Relevant analysis scripts are available in GitHub (https://github.com/jennaswa/arg_bd).

### Authors’ Contributions

JMS led the long-read sequencing analysis and drafted the manuscript. ERF led the 16S rRNA gene amplicon sequencing analysis. JMS, LH, ARH, and MAA performed laboratory procedures. JMS and ERF collected animal fecal samples. ESG and SMS contributed archived human stool samples. JMS, ERF, ABB, and AJP conceived the study. All authors contributed to and approved the final manuscript.

## Acknowledgements

We sincerely thank the study participants as well as the icddr,b staff for assistance with animal feces sample collection. Thank you to Laura Kwong for assisting with fecal composites in Bangladesh. We also thank Rebecca Batorsky, Marlene Wolfe, and Colin Worby for insight on bioinformatic analyses.

