## Supplemental Information for "Differential overlap in human and animal fecal microbiomes and resistomes in rural versus urban Bangladesh"

**Table S1. Quality control parameters for DNA extraction for long-read sequencing. Fragment size distributions were not determined for extraction blanks and are marked "*NA*". For samples with fragment size distributions marked "*NA*", peaks could not be detected in the sample's electropherogram by the Agilent Bioanalyzer.**

| | | Purity | | Fragment Size Distribution | | |
|---|---|---|---|---|---|---|
| **Fecal Host** | **Concentration (ng/uL)** | **OD 260/280** | **OD 260/230** | **From (bp)** | **To (bp)** | **Average (bp)** |
| Rural Chicken 1 | 2.172 | 1.65 | 0.10 | *NA* | *NA* | *NA* |
| Rural Chicken 2 | 2.227 | 1.61 | 0.59 | 683 | 27808 | 5440 |
| Rural Chicken 3 | 1.480 | 1.91 | 0.47 | *NA* | *NA* | *NA* |
| Rural Chicken 4 | 1.146 | 2.08 | 0.30 | *NA* | *NA* | *NA* |
| Urban Chicken 1 | 39.12 | 1.66 | 1.45 | 562 | 28274 | 5521 |
| Urban Chicken 2 | 16.58 | 1.12 | 0.73 | 606 | 28157 | 4454 |
| Urban Chicken 3 | 3.014 | 1.28 | 0.02 | *NA* | *NA* | *NA* |
| Urban Chicken 4 | 4.250 | 1.73 | 0.53 | 521 | 26442 | 4932 |
| Rural Goat 1 | 53.71 | 1.67 | 0.82 | 656 | 29090 | 6103 |
| Rural Goat 2 | 85.33 | 1.76 | 1.22 | 689 | 37014 | 5921 |
| Rural Goat 3 | 121.3 | 1.82 | 1.84 | 491 | 32469 | 5978 |
| Rural Goat 4 | 67.46 | 1.71 | 1.07 | 337 | 42375 | 6059 |
| Urban Goat 1 | 102.5 | 1.83 | 1.81 | 556 | 35499 | 7202 |
| Urban Goat 2 | 96.72 | 1.79 | 1.34 | *NA* | *NA* | *NA* |
| Urban Goat 3 | 220.7 | 1.81 | 1.49 | 30 | 29556 | 5670 |
| Urban Goat 4 | 147.5 | 1.76 | 1.24 | 405 | 30975 | 7748 |
| Rural Human 1 | 120.1 | 1.85 | 1.84 | 620 | 26875 | 5994 |
| Rural Human 2 | 144.7 | 1.87 | 1.92 | 332 | 37713 | 5150 |
| Rural Human 3 | 114.3 | 1.87 | 1.99 | 667 | 31653 | 7051 |
| Rural Human 4 | 55.23 | 1.87 | 1.74 | 665 | 45836 | 7777 |
| Urban Human 1 | 121.6 | 1.87 | 0.82 | 504 | 31770 | 6400 |
| Urban Human 2 | 37.58 | 1.87 | 1.55 | 416 | 35499 | 5727 |
| Urban Human 3 | 40.15 | 1.89 | 1.41 | 570 | 30605 | 5138 |
| Urban Human 4 | 38.32 | 1.86 | 1.62 | *NA* | *NA* | *NA* |
| Extraction Blank 1 | <0.100 | 1.97 | 0.08 | *NA* | *NA* | *NA* |
| Extraction Blank 2 | <0.100 | 1.36 | 0.31 | *NA* | *NA* | *NA* |
| Extraction Blank 3 | <0.100 | 4.97 | 0.07 | *NA* | *NA* | *NA* |

**Table S2. Illumina primers**

| Illumina 5' Adapter | Golay Barcode | Primer Pad | Primer Linker | Primer |
|---|---|---|---|---|
| CAAGCAGAAGACGGCATA CGAGAT[a] | - | AGTCAGCCAG | CC | GGACTACNVGGGTWTCTAA T |
| AATGATACGGCGACCACCG AGATCTACACGCT | AGCCTTCGTCGC | TATGGTAATT | GT | GTGYCAGCMGCCGCGGTAA |
| AATGATACGGCGACCACCG AGATCTACACGCT | TCCATACCGGAA | TATGGTAATT | GT | GTGYCAGCMGCCGCGGTAA |
| AATGATACGGCGACCACCG AGATCTACACGCT | AGCCCTGCTACA | TATGGTAATT | GT | GTGYCAGCMGCCGCGGTAA |
| AATGATACGGCGACCACCG AGATCTACACGCT | CCTAACGGTCCA | TATGGTAATT | GT | GTGYCAGCMGCCGCGGTAA |
| AATGATACGGCGACCACCG AGATCTACACGCT | CGCGCCTTAAAC | TATGGTAATT | GT | GTGYCAGCMGCCGCGGTAA |
| AATGATACGGCGACCACCG AGATCTACACGCT | TATGGTACCCAG | TATGGTAATT | GT | GTGYCAGCMGCCGCGGTAA |
| AATGATACGGCGACCACCG AGATCTACACGCT | TACAATATCTGT | TATGGTAATT | GT | GTGYCAGCMGCCGCGGTAA |
| AATGATACGGCGACCACCG AGATCTACACGCT | AATTTAGGTAGG | TATGGTAATT | GT | GTGYCAGCMGCCGCGGTAA |
| AATGATACGGCGACCACCG AGATCTACACGCT | GACTCAACCAGT | TATGGTAATT | GT | GTGYCAGCMGCCGCGGTAA |
| AATGATACGGCGACCACCG AGATCTACACGCT | GCCTCTACGTCG | TATGGTAATT | GT | GTGYCAGCMGCCGCGGTAA |
| AATGATACGGCGACCACCG AGATCTACACGCT | ACTACTGAGGAT | TATGGTAATT | GT | GTGYCAGCMGCCGCGGTAA |
| AATGATACGGCGACCACCG AGATCTACACGCT | AATTCACCTCCT | TATGGTAATT | GT | GTGYCAGCMGCCGCGGTAA |
| AATGATACGGCGACCACCG AGATCTACACGCT | CGTATAAATGCG | TATGGTAATT | GT | GTGYCAGCMGCCGCGGTAA |
| AATGATACGGCGACCACCG AGATCTACACGCT | ATGCTGCAACAC | TATGGTAATT | GT | GTGYCAGCMGCCGCGGTAA |
| AATGATACGGCGACCACCG AGATCTACACGCT | ACTCGCTCGCTG | TATGGTAATT | GT | GTGYCAGCMGCCGCGGTAA |
| AATGATACGGCGACCACCG AGATCTACACGCT | TTCCTTAGTAGT | TATGGTAATT | GT | GTGYCAGCMGCCGCGGTAA |
| AATGATACGGCGACCACCG AGATCTACACGCT | CGTCCGTATGAA | TATGGTAATT | GT | GTGYCAGCMGCCGCGGTAA |
| AATGATACGGCGACCACCG AGATCTACACGCT | ACGTGAGGAACG | TATGGTAATT | GT | GTGYCAGCMGCCGCGGTAA |
| AATGATACGGCGACCACCG AGATCTACACGCT | GTTGATACGATG | TATGGTAATT | GT | GTGYCAGCMGCCGCGGTAA |
| AATGATACGGCGACCACCG AGATCTACACGCT | GTCAACGCTGTC | TATGGTAATT | GT | GTGYCAGCMGCCGCGGTAA |
| AATGATACGGCGACCACCG AGATCTACACGCT | TGAGACCCTACA | TATGGTAATT | GT | GTGYCAGCMGCCGCGGTAA |
| AATGATACGGCGACCACCG AGATCTACACGCT | ACTTGGTGTAAG | TATGGTAATT | GT | GTGYCAGCMGCCGCGGTAA |
| AATGATACGGCGACCACCG AGATCTACACGCT | ATTACGTATCAT | TATGGTAATT | GT | GTGYCAGCMGCCGCGGTAA |
| AATGATACGGCGACCACCG AGATCTACACGCT | CACGCAGTCTAC | TATGGTAATT | GT | GTGYCAGCMGCCGCGGTAA |

| | | | | |
|---|---|---|---|---|
| AATGATACGGCGACCACCG AGATCTACACGCT | TGTGCACGCCAT | TATGGTAATT | GT | GTGYCAGCMGCCGCGGTAA |
| AATGATACGGCGACCACCG AGATCTACACGCT | CCGGACAAGAAG | TATGGTAATT | GT | GTGYCAGCMGCCGCGGTAA |
| AATGATACGGCGACCACCG AGATCTACACGCT | TTGCTGGACGCT | TATGGTAATT | GT | GTGYCAGCMGCCGCGGTAA |
| AATGATACGGCGACCACCG AGATCTACACGCT | TACTAACGCGGT | TATGGTAATT | GT | GTGYCAGCMGCCGCGGTAA |
| AATGATACGGCGACCACCG AGATCTACACGCT | GCGATCACACCT | TATGGTAATT | GT | GTGYCAGCMGCCGCGGTAA |
| AATGATACGGCGACCACCG AGATCTACACGCT | CAAACGCACTAA | TATGGTAATT | GT | GTGYCAGCMGCCGCGGTAA |
| AATGATACGGCGACCACCG AGATCTACACGCT | GAAGAGGGTTGA | TATGGTAATT | GT | GTGYCAGCMGCCGCGGTAA |
| AATGATACGGCGACCACCG AGATCTACACGCT | TGAGTGGTCTGT | TATGGTAATT | GT | GTGYCAGCMGCCGCGGTAA |
| AATGATACGGCGACCACCG AGATCTACACGCT | TTACACAAAGGC | TATGGTAATT | GT | GTGYCAGCMGCCGCGGTAA |
| AATGATACGGCGACCACCG AGATCTACACGCT | ACGACGCATTTG | TATGGTAATT | GT | GTGYCAGCMGCCGCGGTAA |
| AATGATACGGCGACCACCG AGATCTACACGCT | TATCCAAGCGCA | TATGGTAATT | GT | GTGYCAGCMGCCGCGGTAA |
| AATGATACGGCGACCACCG AGATCTACACGCT | AGAGCCAAGAGC | TATGGTAATT | GT | GTGYCAGCMGCCGCGGTAA |
| AATGATACGGCGACCACCG AGATCTACACGCT | GGTTGCCCTGTA | TATGGTAATT | GT | GTGYCAGCMGCCGCGGTAA |
| AATGATACGGCGACCACCG AGATCTACACGCT | CATATAGCCCGA | TATGGTAATT | GT | GTGYCAGCMGCCGCGGTAA |
| AATGATACGGCGACCACCG AGATCTACACGCT | GCCTATGAGATC | TATGGTAATT | GT | GTGYCAGCMGCCGCGGTAA |
| AATGATACGGCGACCACCG AGATCTACACGCT | CAAGTGAAGGGA | TATGGTAATT | GT | GTGYCAGCMGCCGCGGTAA |
| AATGATACGGCGACCACCG AGATCTACACGCT | CACGTTTATTCC | TATGGTAATT | GT | GTGYCAGCMGCCGCGGTAA |
| AATGATACGGCGACCACCG AGATCTACACGCT | TAATCGGTGCCA | TATGGTAATT | GT | GTGYCAGCMGCCGCGGTAA |
| AATGATACGGCGACCACCG AGATCTACACGCT | TGACTAATGGCC | TATGGTAATT | GT | GTGYCAGCMGCCGCGGTAA |
| AATGATACGGCGACCACCG AGATCTACACGCT | CGGGACACCCGA | TATGGTAATT | GT | GTGYCAGCMGCCGCGGTAA |
| AATGATACGGCGACCACCG AGATCTACACGCT | CTGTCTATACTA | TATGGTAATT | GT | GTGYCAGCMGCCGCGGTAA |
| AATGATACGGCGACCACCG AGATCTACACGCT | TATGCCAGAGAT | TATGGTAATT | GT | GTGYCAGCMGCCGCGGTAA |
| AATGATACGGCGACCACCG AGATCTACACGCT | CGTTTGGAATGA | TATGGTAATT | GT | GTGYCAGCMGCCGCGGTAA |
| AATGATACGGCGACCACCG AGATCTACACGCT | AAGAACTCATGA | TATGGTAATT | GT | GTGYCAGCMGCCGCGGTAA |
| AATGATACGGCGACCACCG AGATCTACACGCT | TGATATCGTCTT | TATGGTAATT | GT | GTGYCAGCMGCCGCGGTAA |
| AATGATACGGCGACCACCG AGATCTACACGCT | CGGTGACCTACT | TATGGTAATT | GT | GTGYCAGCMGCCGCGGTAA |
| AATGATACGGCGACCACCG AGATCTACACGCT | AATGCGCGTATA | TATGGTAATT | GT | GTGYCAGCMGCCGCGGTAA |

| | | | | |
|---|---|---|---|---|
| AATGATACGGCGACCACCG AGATCTACACGCT | CTTGATTCTTGA | TATGGTAATT | GT | GTGYCAGCMGCCGCGGTAA |
| AATGATACGGCGACCACCG AGATCTACACGCT | GAAATCTTGAAG | TATGGTAATT | GT | GTGYCAGCMGCCGCGGTAA |
| AATGATACGGCGACCACCG AGATCTACACGCT | GAGATACAGTTC | TATGGTAATT | GT | GTGYCAGCMGCCGCGGTAA |
| AATGATACGGCGACCACCG AGATCTACACGCT | GGTGAGCAAGCA | TATGGTAATT | GT | GTGYCAGCMGCCGCGGTAA |
| AATGATACGGCGACCACCG AGATCTACACGCT | TAAATATACCCT | TATGGTAATT | GT | GTGYCAGCMGCCGCGGTAA |
| AATGATACGGCGACCACCG AGATCTACACGCT | TTGCGGACCCTA | TATGGTAATT | GT | GTGYCAGCMGCCGCGGTAA |
| AATGATACGGCGACCACCG AGATCTACACGCT | GTCGTCCAAATG | TATGGTAATT | GT | GTGYCAGCMGCCGCGGTAA |
| AATGATACGGCGACCACCG AGATCTACACGCT | TGCACAGTCGCT | TATGGTAATT | GT | GTGYCAGCMGCCGCGGTAA |
| AATGATACGGCGACCACCG AGATCTACACGCT | TTACTGTGGCCG | TATGGTAATT | GT | GTGYCAGCMGCCGCGGTAA |
| AATGATACGGCGACCACCG AGATCTACACGCT | GGTTCATGAACA | TATGGTAATT | GT | GTGYCAGCMGCCGCGGTAA |
| AATGATACGGCGACCACCG AGATCTACACGCT | TAACAATAATTC | TATGGTAATT | GT | GTGYCAGCMGCCGCGGTAA |
| AATGATACGGCGACCACCG AGATCTACACGCT | CTTATTAAACGT | TATGGTAATT | GT | GTGYCAGCMGCCGCGGTAA |
| AATGATACGGCGACCACCG AGATCTACACGCT | GCTCGAAGATTC | TATGGTAATT | GT | GTGYCAGCMGCCGCGGTAA |
| AATGATACGGCGACCACCG AGATCTACACGCT | TATTTGATTGGT | TATGGTAATT | GT | GTGYCAGCMGCCGCGGTAA |
| AATGATACGGCGACCACCG AGATCTACACGCT | TGTCAAAGTGAC | TATGGTAATT | GT | GTGYCAGCMGCCGCGGTAA |
| AATGATACGGCGACCACCG AGATCTACACGCT | CTATGTATTAGT | TATGGTAATT | GT | GTGYCAGCMGCCGCGGTAA |
| AATGATACGGCGACCACCG AGATCTACACGCT | ACTCCCGTGTGA | TATGGTAATT | GT | GTGYCAGCMGCCGCGGTAA |
| AATGATACGGCGACCACCG AGATCTACACGCT | CGGTATAGCAAT | TATGGTAATT | GT | GTGYCAGCMGCCGCGGTAA |
| AATGATACGGCGACCACCG AGATCTACACGCT | GACTCTGCTCAG | TATGGTAATT | GT | GTGYCAGCMGCCGCGGTAA |
| AATGATACGGCGACCACCG AGATCTACACGCT | GTCATGCTCCAG | TATGGTAATT | GT | GTGYCAGCMGCCGCGGTAA |
| AATGATACGGCGACCACCG AGATCTACACGCT | TACCGAAGGTAT | TATGGTAATT | GT | GTGYCAGCMGCCGCGGTAA |
| AATGATACGGCGACCACCG AGATCTACACGCT | GTGGAGTCTCAT | TATGGTAATT | GT | GTGYCAGCMGCCGCGGTAA |
| AATGATACGGCGACCACCG AGATCTACACGCT | ACCTTACACCTT | TATGGTAATT | GT | GTGYCAGCMGCCGCGGTAA |
| AATGATACGGCGACCACCG AGATCTACACGCT | TAATCTCGCCGG | TATGGTAATT | GT | GTGYCAGCMGCCGCGGTAA |
| AATGATACGGCGACCACCG AGATCTACACGCT | ATCTAGTGGCAA | TATGGTAATT | GT | GTGYCAGCMGCCGCGGTAA |
| AATGATACGGCGACCACCG AGATCTACACGCT | ACGCTTAACGAC | TATGGTAATT | GT | GTGYCAGCMGCCGCGGTAA |
| AATGATACGGCGACCACCG AGATCTACACGCT | TACGGATTATGG | TATGGTAATT | GT | GTGYCAGCMGCCGCGGTAA |

| | | | | |
|---|---|---|---|---|
| AATGATACGGCGACCACCG AGATCTACACGCT | ATACATGCAAGA | TATGGTAATT | GT | GTGYCAGCMGCCGCGGTAA |
| AATGATACGGCGACCACCG AGATCTACACGCT | CTTAGTGCAGAA | TATGGTAATT | GT | GTGYCAGCMGCCGCGGTAA |
| AATGATACGGCGACCACCG AGATCTACACGCT | AATCTTGCGCCG | TATGGTAATT | GT | GTGYCAGCMGCCGCGGTAA |
| AATGATACGGCGACCACCG AGATCTACACGCT | AGGATCAGGGAA | TATGGTAATT | GT | GTGYCAGCMGCCGCGGTAA |
| AATGATACGGCGACCACCG AGATCTACACGCT | AATAACTAGGGT | TATGGTAATT | GT | GTGYCAGCMGCCGCGGTAA |
| AATGATACGGCGACCACCG AGATCTACACGCT | TATTGCAGCAGC | TATGGTAATT | GT | GTGYCAGCMGCCGCGGTAA |
| AATGATACGGCGACCACCG AGATCTACACGCT | TGATGTGCTAAG | TATGGTAATT | GT | GTGYCAGCMGCCGCGGTAA |
| AATGATACGGCGACCACCG AGATCTACACGCT | GTAGTAGACCAT | TATGGTAATT | GT | GTGYCAGCMGCCGCGGTAA |
| AATGATACGGCGACCACCG AGATCTACACGCT | AGTAAAGATCGT | TATGGTAATT | GT | GTGYCAGCMGCCGCGGTAA |
| AATGATACGGCGACCACCG AGATCTACACGCT | CTCGCCCTCGCC | TATGGTAATT | GT | GTGYCAGCMGCCGCGGTAA |
| AATGATACGGCGACCACCG AGATCTACACGCT | TCTCTTTCGACA | TATGGTAATT | GT | GTGYCAGCMGCCGCGGTAA |
| AATGATACGGCGACCACCG AGATCTACACGCT | ACATACTGAGCA | TATGGTAATT | GT | GTGYCAGCMGCCGCGGTAA |
| AATGATACGGCGACCACCG AGATCTACACGCT | TGAGTATGAGTA | TATGGTAATT | GT | GTGYCAGCMGCCGCGGTAA |
| AATGATACGGCGACCACCG AGATCTACACGCT | AATGGTTCAGCA | TATGGTAATT | GT | GTGYCAGCMGCCGCGGTAA |
| AATGATACGGCGACCACCG AGATCTACACGCT | GAACCAGTACTC | TATGGTAATT | GT | GTGYCAGCMGCCGCGGTAA |
| AATGATACGGCGACCACCG AGATCTACACGCT | CGCACCCATACA | TATGGTAATT | GT | GTGYCAGCMGCCGCGGTAA |
| AATGATACGGCGACCACCG AGATCTACACGCT | GTGCCATAATCG | TATGGTAATT | GT | GTGYCAGCMGCCGCGGTAA |
| AATGATACGGCGACCACCG AGATCTACACGCT | ACTCTTACTTAG | TATGGTAATT | GT | GTGYCAGCMGCCGCGGTAA |

[a] This is the reverse primer used. The Illumina adapter is the reverse complement, the pad is the reverse pad, the linker is the reverse linker, and the primer is the 806rB primer. All others listed are the barcoded forward primers with the 515f primer.

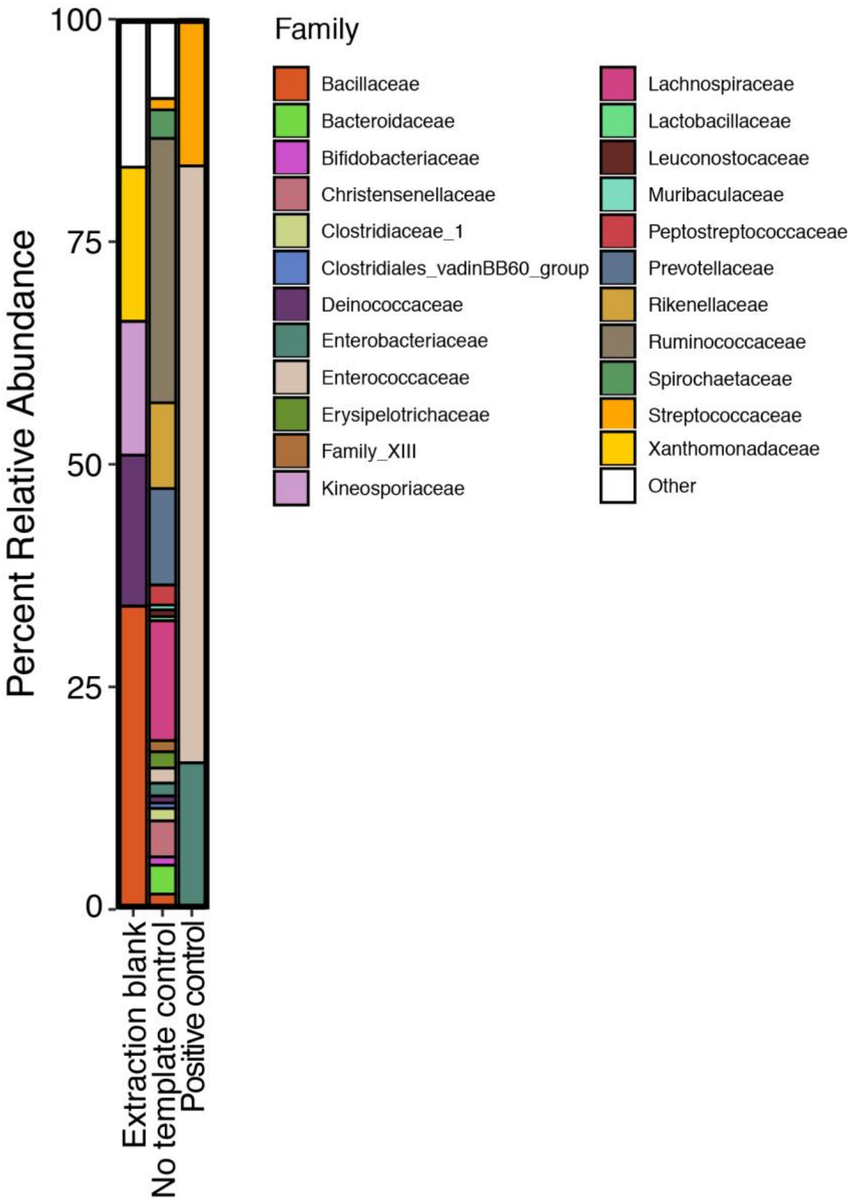

**Figure S1. Relative abundance of the 30 most abundant families in all controls from 16S rRNA gene analysis. All other taxa are grouped into "Other".**

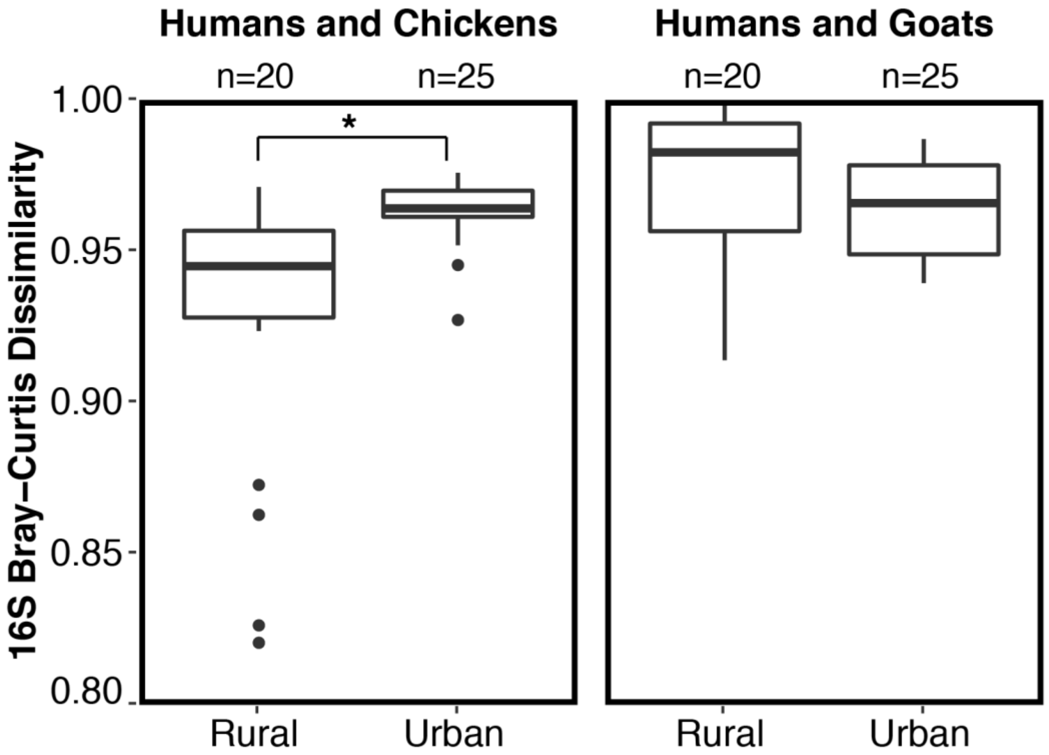

**Figure S2. Bray-Curtis dissimilarity between bacterial community compositions in humans and chickens (left) and in humans and goats (right) with taxa present in no template control removed**

**Table S3. Descriptive statistics for long-read sequencing and assembly**

| Fecal Host | Raw Reads | | | | Assembled Contigs | | |
|---|---|---|---|---|---|---|---|
| | Number Reads (millions) | Mean Length [SD] (bp) | Total Data (Gbp) | Bacterial Data (Gbp) | Number Contigs (thousands) | N50 (kbp) | Median Length (kbp) |
| Rural Chicken | 6.415 | 668.8 [1013] | 4.291 | 2.747 | 1.636 | 34.33 | 21.79 |
| Urban Chicken | 9.834 | 809.2 [1151] | 7.958 | 5.129 | 4.598 | 44.95 | 21.54 |
| Rural Goat | 11.19 | 2096 [2699] | 23.44 | 14.98 | 38.60 | 44.73 | 24.15 |
| Urban Goat | 8.821 | 2391 [3367] | 21.09 | 14.77 | 24.52 | 60.33 | 31.62 |
| Rural Human | 6.610 | 2864 [2799] | 18.93 | 14.22 | 17.73 | 60.71 | 22.82 |
| Urban Human | 9.964 | 2327 [2406] | 23.18 | 18.05 | 27.00 | 53.86 | 22.03 |

**Table S4. Comparisons of normalized abundance of antimicrobial resistance genes in humans, chickens, and goats using Wilcoxon rank sum tests**

| | | Chicken | | Goat | | Human | | | |
|---|---|---|---|---|---|---|---|---|---|
| **Comparison** | **Location** | ***N*** | **Rank Sum** | ***N*** | **Rank Sum** | ***N*** | **Rank Sum** | ***Z*-score** | ***p*-Value** |
| Chicken vs. Human | Rural | 4 | 21 | *NA* | *NA* | 4 | 15 | 0.866 | 0.387 |
| | Urban | 4 | 21 | *NA* | *NA* | 4 | 15 | 0.866 | 0.387 |
| Goat vs. Human | Rural | *NA* | *NA* | 3 | 6 | 4 | 22 | -2.121 | 0.034 |
| | Urban | *NA* | *NA* | 4 | 10 | 4 | 26 | -2.309 | 0.021 |
| Chicken vs. Goat | Rural | 4 | 26 | 3 | 10 | *NA* | *NA* | 2.121 | 0.034 |
| | Urban | 4 | 26 | 4 | 10 | *NA* | *NA* | 2.309 | 0.021 |

**Table S5. Comparisons of normalized abundance of antimicrobial resistance genes, paired by drug class, in rural versus urban communities using Wilcoxon signed-rank tests**

| **Fecal Host** | ***N*** | ***Z*-score** | ***p*-Value** |
|---|---|---|---|
| Chicken | 13 | -1.992 | 0.046 |
| Goat | 13 | -2.385 | 0.017 |
| Human | 13 | -1.051 | 0.293 |

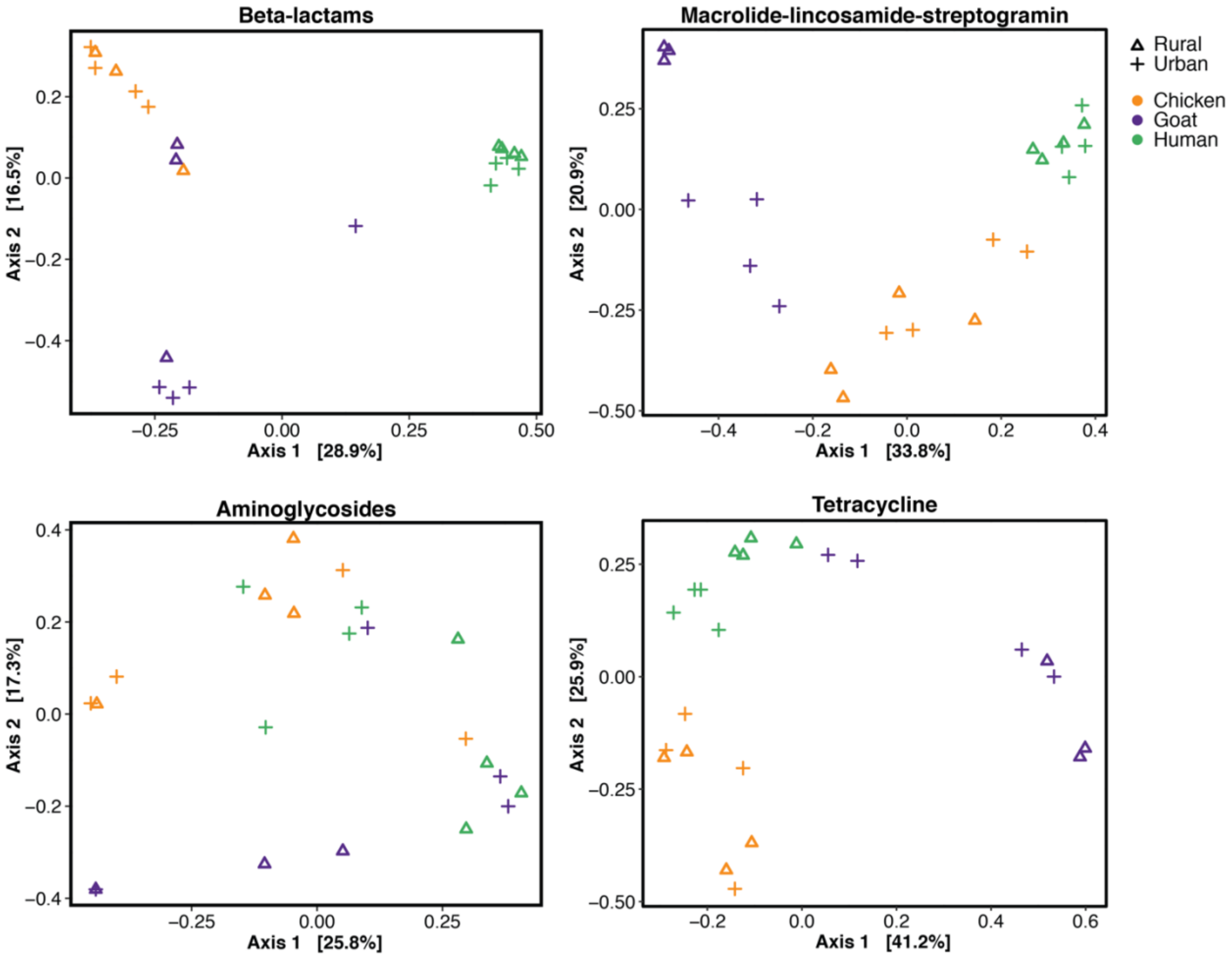

**Figure S3. Principal coordinate analyses (PCoAs) of Bray-Curtis dissimilarity between antimicrobial resistance gene alleles in humans, chickens, and goats for four most abundant drug classes. Color indicates fecal host and symbol indicates rural, urban, or control.**

**Table S6. BLASTN confirmation of potential pathogens classified to the strain or subspecies level and carrying antimicrobial resistance genes**

| Fecal Host | Potential Pathogen | NCBI Tax ID | Accession | Maximum Score | Total Score | Query Coverage | E-Value | Percent Identity |
|---|---|---|---|---|---|---|---|---|
| Rural Human | *Escherichia coli PCN033* | 1001989 | CP006632.1 | 33968 | 2.61E+05 | 77% | 0 | 98.32% |
| Urban Human | *Clostridioides difficile M120* | 699035 | FN665653.1 | 5694 | 5694 | 9% | 0 | 99.49% |
| Urban Goat | *Clostridioides difficile M120* | 699035 | FN665653.1 | 3179 | 3442 | 7% | 0 | 98.83% |
